# Pneumococcal colonization impairs nasal and lung mucosal immune responses to Live Attenuated Influenza Vaccination in adults

**DOI:** 10.1101/2020.02.24.20025098

**Authors:** Beatriz F. Carniel, Fernando Marcon, Jamie Rylance, Seher Zaidi, Jesus Reine, Edessa Negera, Elissavet Nikolaou, Sherin Pojar, Carla Solórzano, Andrea Collins, Victoria Connor, Debby Bogaert, Stephen B. Gordon, Helder Nakaya, Daniela M. Ferreira, Simon P. Jochems, Elena Mitsi

**Affiliations:** Department of Clinical Sciences, Liverpool School of Tropical Medicine, Liverpool, United Kingdom; Royal Liverpool and Broadgreen University Hospital, Liverpool, United Kingdom; Centre for Inflammation Research, University of Edinburgh, Edinburgh, United Kingdom; Department of Paediatric Immunology and Infectious Diseases, Utrecht, The Netherlands; Malawi Liverpool Wellcome Trust Clinical Research Programme, College of Medicine, Blantyre, Malawi; Department of Clinical and Toxicological Analyses, School of Pharmaceutical Sciences, University of São Paulo, São Paolo, Brazil

**Keywords:** Live attenuated influenza vaccine, *S. pneumoniae*, pneumococcal colonization, cellular responses, human mucosal sites

## Abstract

Influenza virus infections affect millions of people annually. Current available vaccines provide varying rates of protection. There is a knowledge gap on how the nasal microbiota, particularly established pneumococcal colonization, shapes the response to influenza vaccination. In this study, we inoculated healthy adults with live *S. pneumoniae* and vaccinated them three days later with either TIV or LAIV. Vaccine-induced immune responses were assessed in nose, blood and lung. Nasal pneumococcal colonization had no impact upon TIV-induced antibody responses to influenza, which manifested in all compartments. However, pre-existing pneumococcal colonization dampened LAIV-mediated mucosal antibody responses, primarily IgA in the nose and IgG in the lung. Pulmonary influenza-specific cellular responses were more apparent in the LAIV group compared to either TIV or an unvaccinated group. These results indicate that TIV and LAIV elicit differential immunity to adults and that LAIV immunogenicity is diminished by the nasal presence of *S. pneumoniae*. This important confounder should be considered when assessing LAIV efficacy.

## INTRODUCTION

Each year, 5–15% of the world’s population will suffer from an influenza infection, with up to 5 million cases of severe disease and 500,000 deaths^1^. Influenza viruses have the ability to mutate and hence escape immune defence mechanisms, necessitating annual vaccine updates. These vaccines include the tetravalent inactivated influenza vaccine (TIV)^2^, which is given intramuscularly, and the live attenuated influenza vaccine (LAIV)^3^, which is administered intranasally. The route of vaccination can trigger distinct immune mechanisms and pathways of protection. For example, TIV is given as an intramuscular injection and induces neutralizing antibodies to strain-specific glycoproteins hemagglutinin (HA) and neuraminidase (NA)^4^. By comparison, LAIV is intranasally administered as a cold-adapted vaccine that replicates only in the nasopharynx and mimics natural infection^5^. Nasal replication leads to recognition of its pathogen-associated molecular patterns (PAMPs) by host pattern recognition receptors (PRRs), which initiates a cascade of cellular immune responses^6^. In mice, LAIV vaccination increases the frequency of CD4^+^ and CD8^+^ T cells in the lung and cytokine production upon influenza re-stimulation compared to vaccination with the inactivated virus or no vaccine administration^7, 8, 9, 10^. Moreover, LAIV seeds the murine lung with both CD4^+^ tissue resident memory (TRM) and virus-specific CD8^+^ T cells. TRM have been shown to provide long-term cross-strain protection against influenza^7^. In humans, the immune responses elicited by LAIV have been found to provide broader clinical protection in children compared to the inactivated influenza vaccines (IIVs)^11^. However, the detailed immunological mechanisms of this remain incompletely understood.

Influenza vaccines are re-formulated annually to represent circulating strains, but genomic changes over time (antigenic drift) reduce effectiveness^12^. Estimates from the World Health Organization (WHO) suggest that influenza vaccines effectiveness rarely exceeds 60% and has fallen below 30% in some years ^13, 14^. Poor effectiveness of LAIV among 2 to 17-year-olds in 2014 and 2015 led to the Centre for Disease Control (CDC) recommending its temporary exclusion from the US national childhood influenza immunization programme during the subsequent two seasons^15^. From 2018, no such recommendations have been made. Many underlying causes for this variation have been suggested, including: poor matching with circulating strains^16^, differential ability of some LAIV types to induce immunity (in particular against H_1_N_1_ strains^15^), and the microbial community composition at times of LAIV administration^17^.

Despite several reports about the microbiota and its impact on vaccination responses^18, 19, 20, 21^, including responses to influenza vaccine^20, 22^, it remains unclear how the microbiome affects LAIV immunogenicity. In murine models, a prior exposure to *S. pneumoniae* (Spn) alters the anti-viral B cell responses during co-infection with wild-type influenza virus, potentially compromising long-term antiviral antibody-mediated immunity^23^. Colonization of the nasopharynx with pneumococcus is very common during childhood, with a point prevalence of 50% of infants in resource-rich settings and up to 90% in low and middle income countries^24^. A significant interaction between Spn colonisation and influenza vaccination could profoundly impact the utility of vaccination, especially amongst the poorest groups of the world.

We used an experimental human pneumococcal challenge (EHPC) model^25^ to experimentally colonize adults with pneumococcus, who three days later received either LAIV (nasal) or TIV (intramuscular). We showed that in humans, LAIV elicits immune responses primarily at mucosal sites-the nose and lung. Interestingly, pre-existing nasal pneumococcal colonization impacted on LAIV immunogenicity, dampening the LAIV-mediated nasal and lung immune responses.

## RESULTS

We conducted a double-blind randomised controlled clinical trial^26^ in which healthy adults (18 to 48 years of age) were vaccinated with either TIV (n=90) or LAIV (n=80) three days after intranasal challenge with live *S. pneumoniae* (Supplementary Fig. 1). To assess and compare the immune responses elicited by influenza vaccination, we collected a series of samples. Mucosal samples, including nasal wash, nasal scrapes (epithelial and immune cells), nasal lining fluid and bronchoalveolar lavage (BAL), as well as serum samples, were collected from the two experimental groups and stratified according to vaccination and pneumococcal carriage status: 1) TIV vaccinated non-Spn colonized (TIV/Spn-, n= 21), 2) TIV vaccinated Spn-colonized (TIV/Spn+, n= 19), 3) LAIV vaccinated non-Spn colonized (LAIV/Spn-, n= 37) and 4) LAIV vaccinated Spn-colonized (LAIV/Spn+, n= 43). For the assessment of lung immune responses, we included a non-vaccinated cohort as control (n=20, 10 Spn- and 10 Spn+), since we were only able to sample the human lung post challenge/vaccination and not at baseline.

### Spn colonization prevents an acute nasal pro-inflammatory response upon LAIV administration

Vaccine-induced inflammatory responses in the nasal mucosa were assessed by measuring levels of 30 cytokines in the nasal fluid at baseline, day −1 (2 days post Spn challenge but a day before vaccination) and at days 3, 6 and 24 post vaccination. LAIV administration induced a mild pro-inflammatory response, which resembled TIV based on similarity analysis (Figure 1A). In particular, only IP-10 and TNF-α were significantly increased (p < 0.05 by Wilcoxon test with Benjamini-Hochberg for multiple testing correction) at 3 days post LAIV. At day 6, levels of TNF-α remained increased compared to baseline (pre-challenge), and levels of 4 more cytokines (IL-1b, IL12, IL15 and IL-2R) had a transient induction at this time-point (Figure 1B). No other cytokine was significantly induced in either LAIV or TIV group at any timepoint.

**Figure 1.**
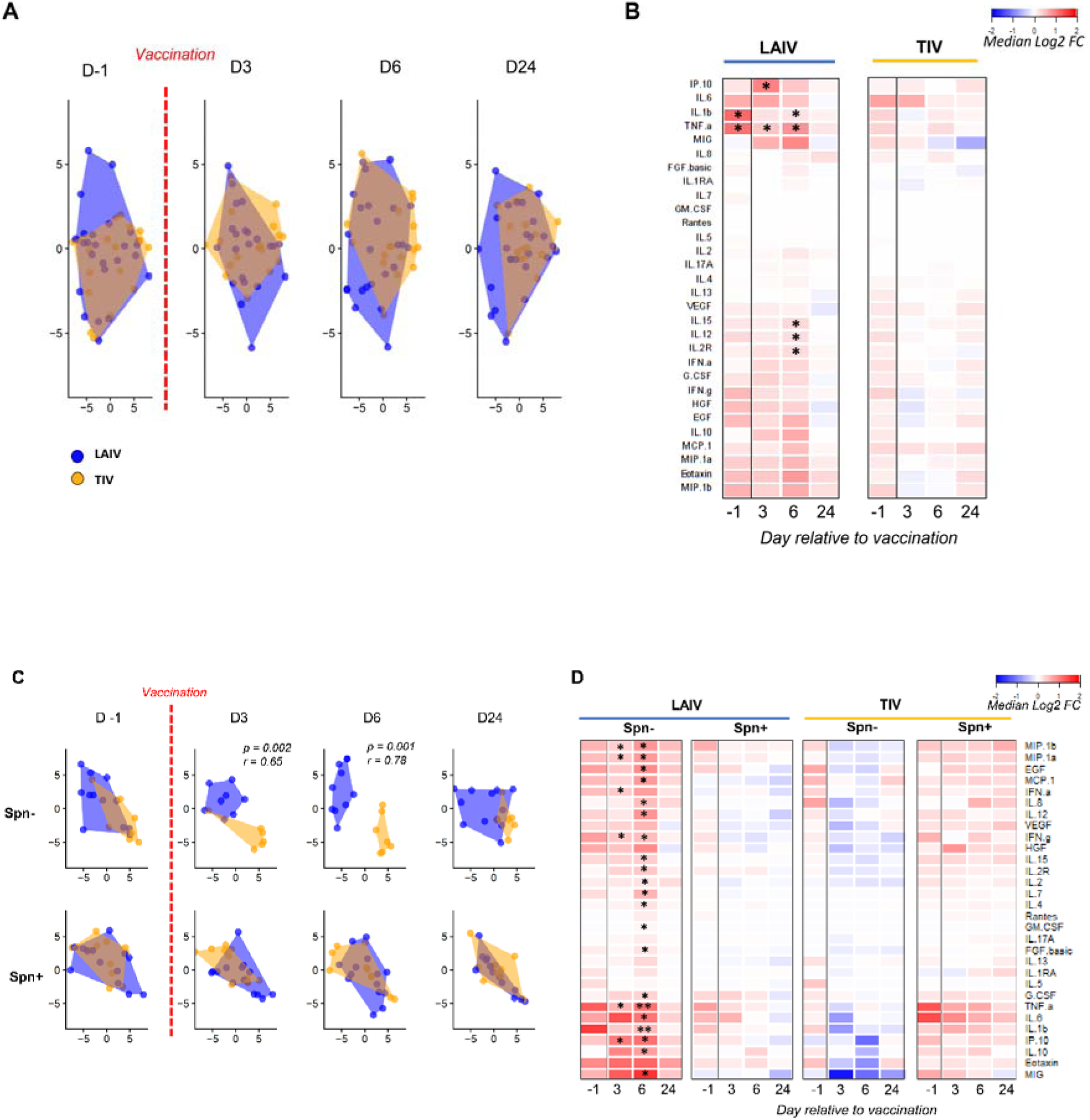
Spn colonization prevents an acute nasal LAIV-induced pro-inflammatory response. Levels of 30 cytokines were measured in nasal fluid at baseline, 1 day before vaccination (D-1), and 3, 6 and 24 days after vaccination for LAIV Spn- (LAIV vaccinated/non-colonized, n=15), LAIV Spn+ (LAIV vaccinated/colonized, n=15), TIV Spn- (TIV vaccinated/non-colonized, n=16) and TIV Spn+ (TIV vaccinated/colonized, n=14). **(A&C)** Samples were clustered based on fold-change levels to baseline using t-distributed stochastic neighbour embedding (t-sne) for LAIV (blue) or TIV (orange). R and P values shown for significant time points based on analysis of similarity (anosim) including fold-changes for all 30 cytokines. **(B)** Heatmap showing median log2 fold change (log2FC) to baseline levels at each of the 4 timepoints after LAIV or TIV administration, with red indicating upregulation and blue downregulation. **(D)** Heatmap showing median log2FC to baseline levels at each of the 4 timepoints for the 4 experimental groups, based on stratification by vaccine and colonization status.**p < 0.01, *p < 0.05 by Wilcoxon paired test, followed by Benjamini-Hochberg correction for multiple testing.

To investigate whether colonization of the nasopharynx with *S. pneumoniae* prior to transient LAIV infection would alter the LAIV-mediated immunogenicity, we stratified the groups according to volunteers’ colonization status and assessed the cytokines profile in the four experimental groups. LAIV induced a transient but robust pro-inflammatory response only in the absence of nasal pneumococcal colonization (Figure 1C). In particular, MIP-1α, MIP-1β, IFN-γ, IFN-α, IP-10 and TNF-α were significantly increased at 3 days post LAIV in the non-colonized group (Figure 1D). At 6 days post LAIV, 21 out of 30 measured cytokines were significantly increased in this group (Figure 1D). No other cytokine was significantly induced in any of the 4 groups at any timepoint.

### LAIV increases the frequency of influenza-specific TNF-α and IFN-γ producing CD4^+^ and TRM CD4^+^ T cells in the lung

Data from animal models suggest that LAIV, but not TIV, induces protective cellular responses in the lung^27, 28^. To assess influenza vaccination-induced cellular responses in the human lung, bronchoalveolar lavage (BAL) cells were stimulated with influenza antigens. T cell subsets (CD4^+^, CD8^+^ and TCR-γδ^+^) were immunophenotyped and cytokine production was measured by intracellular cytokine staining in order to determine the frequency of IFN-γ, IL-17A and TNF-α-producing, influenza-specific T cells (Supplementary Fig. 2). Frequencies of total CD4^+^, CD8^+^ and TCR-γδ^+^ T cells were not affected by vaccination status (Supplementary Fig. 3). Furthermore, we investigated the presence of tissue-resident memory T cell responses to influenza, using the extracellular markers CD69, CD103 and CD49a. As over 1/3 of CD4^+^ CD69^+^ cells, commonly defined as TRM^29^, did not express the additional resident memory markers CD103 and CD49a, we defined TRM only as CD69^+^ (Supplementary Fig. 4). In contrast, nearly all CD8^+^CD69^+^ cells also expressed CD103 and CD49a.

CD4^+^ TNF-α production upon influenza stimulation was observed in both TIV and LAIV recipients regardless of colonization status, but not in unvaccinated individuals, (Figure 2A-B). However, levels of influenza-specific TNF-α were significantly increased in the LAIV/Spn-group compared to the unvaccinated group (median 2.6-fold increase, p = 0.015) (Figure 2B).

**Figure 2.**
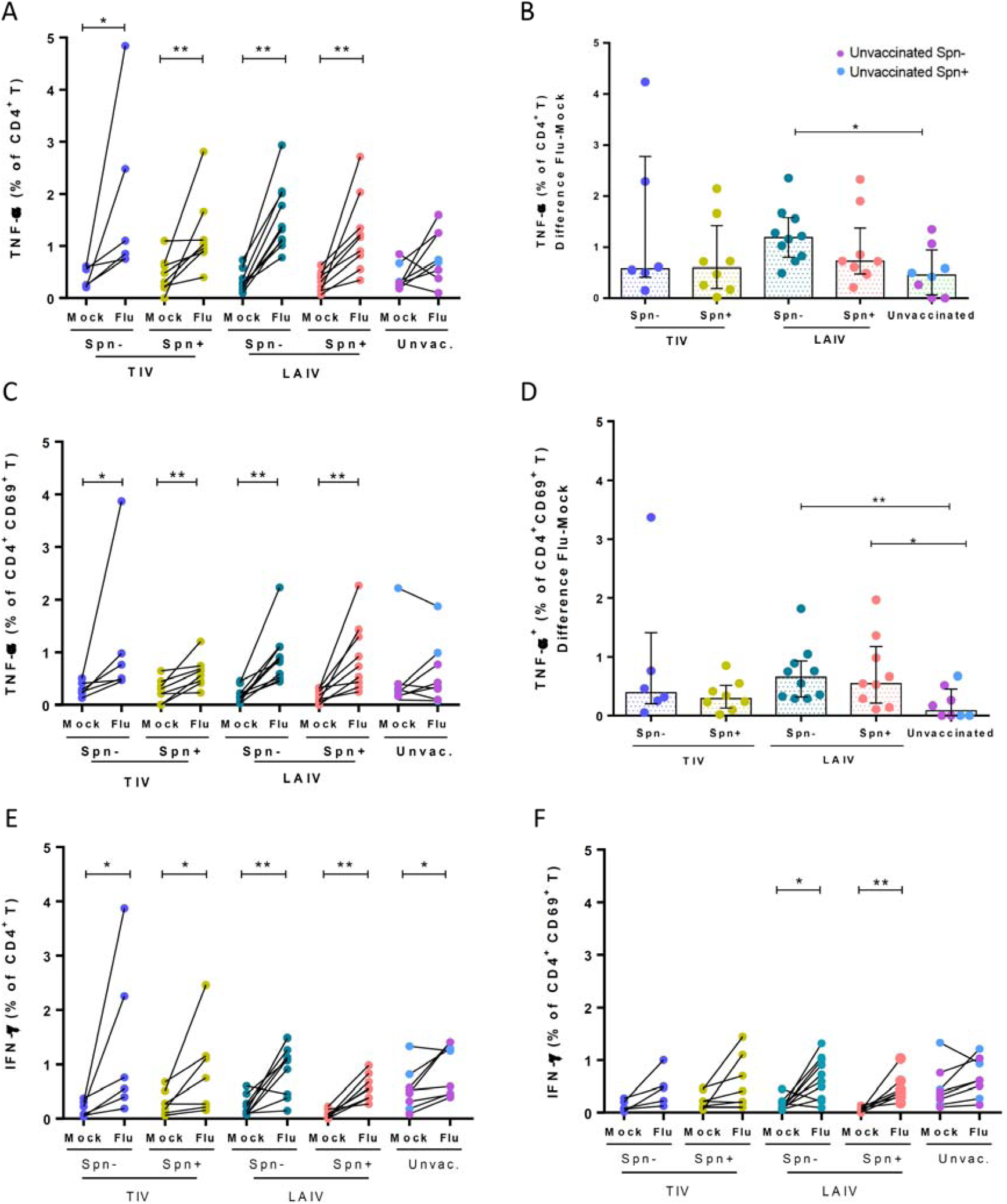
LAIV increases frequency of influenza-specific TNF-α and IFN-γ-producing CD4^+^ and tissue resident memory (TRM) CD4^+^ T cells in the lung. Frequencies of cytokine-producing CD4+ and TRM CD4+ T cells were measured in human BAL samples by intracellular staining flow cytometry analysis with and without (mock) *in vitro* influenza antigen stimulation. Volunteers were divided by vaccine and colonization status in TIV/Spn- (n=6), TIV/Spn+ (n=8), LAIV/Spn- (n=10), LAIV/Spn+ (n=9), unvaccinated (n=8, 3 Spn- and 5 Spn+) group. **(A)** Production of TNF-α by total CD4^+^ T cells in each group [paired unstimulated (mock) and stimulated condition (flu)]. **(B)** influenza-specific production of TNF- α by total CD4^+^ T cells (Difference between influenza-stimulated and unstimulated) in each group. **(C)** Production of TNF-α by CD4^+^ CD69^+^ T-cells in each group. **(D)** Production of influenza-specific TNF-α by CD4^+^ CD69^+^ T-cells in each group. **(E)** Production of IFN- □ by total CD4^+^ T-cells and **(F)** CD4^+^ CD69^+^ T-cells in each group. Each individual dot represents a single volunteer and the conditions from one individual are connected. Medians with IQR are depicted for influenza-specific responses (panels B and D). *p < 0.05, **p < 0.01 by Wilcoxon test for comparisons within the same group and by Mann-Whitney test for between-group comparisons.

Following stimulation with influenza antigens, CD4^+^ TRM T cells produced TNF-α in all vaccinated groups but not in the unvaccinated group (Figure 2C). The induction of TNF-α producing CD4^+^ TRM following stimulation did not significantly differ between TIV and LAIV, but it was more pronounced in the LAIV group, in both the Spn- and Spn+ groups compared to the unvaccinated group (7.7-fold change, p=0.004 and 6.5-fold change, p=0.024 to unvaccinated, respectively) (Figure 2D).

We also assessed IFN-γ production by total CD4^+^ and TRM CD4^+^ T cells residing in the human lung. IFN-γ production by total CD4^+^ T cells was observed in all groups upon stimulation, including the unvaccinated group (Figure 2E). The levels of IFN-γ producing CD4^+^ T cells were not different when comparing vaccinated and unvaccinated groups. The induction of IFN-γ producing CD4^+^ TRM T cells, however, was greater in the LAIV vaccinated group (Figure 2F). In contrast to total CD4^+^ T cells, stimulation of TRMs of unvaccinated individuals did not elicit an IFN-γ response (Figure 2F).

Furthermore, the proportion of IL-17A producing CD4^+^ T cells or CD4^+^ TRM T cells was not affected by vaccination with either TIV or LAIV (Supplementary Fig. 5A-B).

### LAIV increases the frequency of influenza-specific TNF-α producing CD8^+^ and TRM CD8^+^ T cells in the lungs

*In vitro* re-stimulation with influenza induced increased production of TNF-α by CD8^+^ T cells in LAIV but not TIV or unvaccinated group (Figure 3). When volunteers were stratified based on colonisation status, LAIV/Spn- had a median 2.3-fold increase of TNF-α producing CD8^+^T cells post stimulation compared to the non-stimulated condition (p=0.03). LAIV/Spn+ group had a similar induction on this type of cellular response (median 1.9-fold increase, p=0.007) (Figure 3A). Similarly, TNF-α production by TRM CD8^+^ cells was only observed in the LAIV-vaccinated group, increased by median 3.1- (p=0.006) and 2.1- (p=0.004) fold change in LAIV/Spn- and LAIV Spn+ group, respectively (Figure 3B).

**Figure 3.**
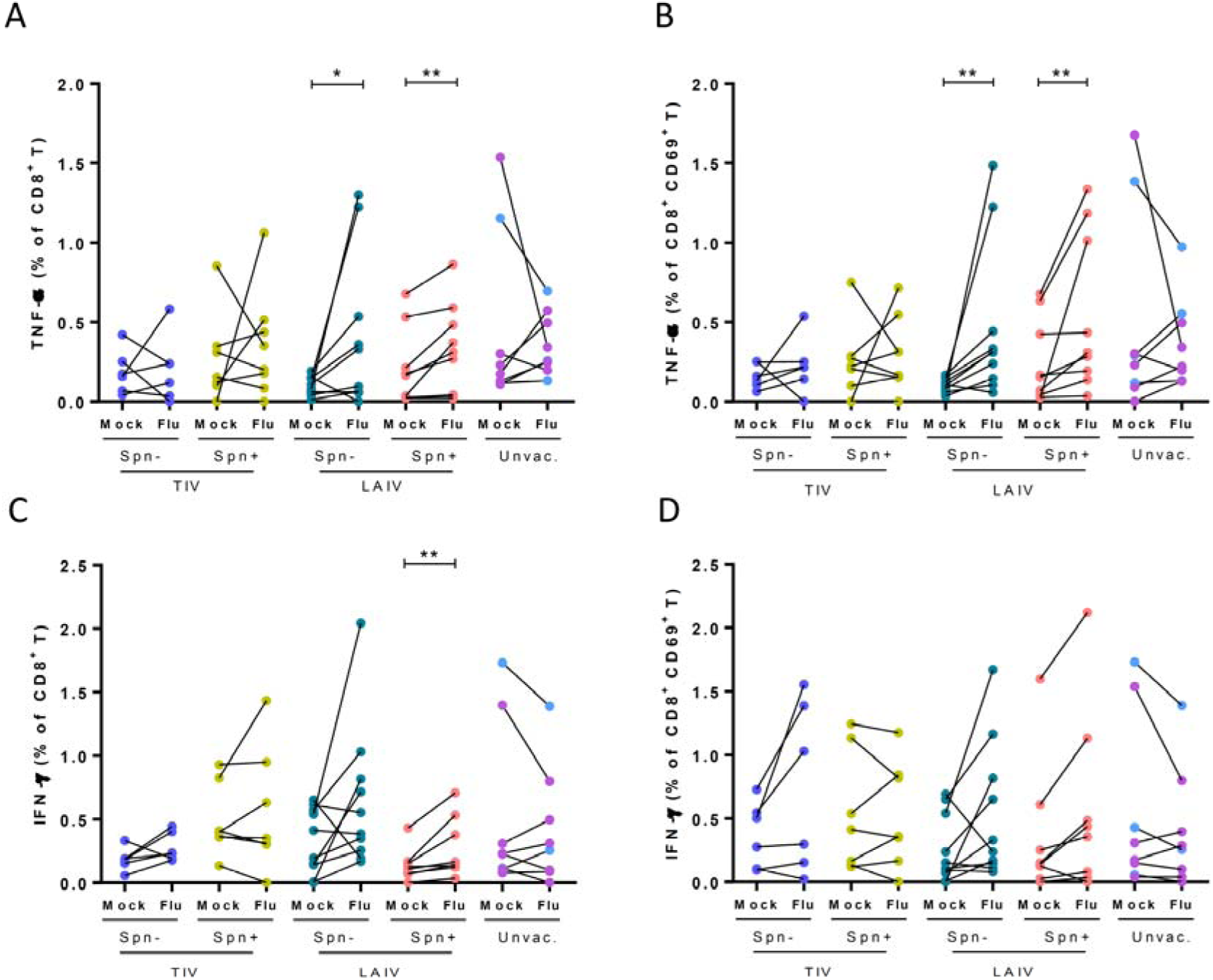
LAIV increases frequency of influenza-specific TNF-α producing CD8^+^ and tissue-resident memory CD8^+^ T cells in the lungs. Frequencies of cytokine-producing CD8^+^ T cells were measured in human BAL samples by intracellular staining flow cytometry analysis following stimulation with influenza antigens or non-stimulation (mock) in each group. Volunteers were divided by vaccine and colonization status in TIV/Spn- (n=6), TIV/Spn- (n=8), LAIV/Spn- (n=10), LAIV/Spn+ (n=9) and unvaccinated (n=8, 3 Spn- and 5 Spn+) group. Production of TNF-α by **(A)** total CD8^+^ T-cells and **(B)** TRM CD8^+^ T-cells in each group (paired unstimulated [mock] and stimulated condition [Flu]). Production of IFN- □ production by **(C)** total CD8^+^ T-cells and **(D)** TRM CD8^+^ T-cells in each group. Each individual dot represents a single volunteer and the conditions per individual are connected. *p < 0.05, **p < 0.01 by Wilcoxon test.

IFN-γ responses by lung CD8^+^ T cells post stimulation were confined in the LAIV group. Although, both LAIV/Spn- and LAIV/Spn+ groups had the same levels of induction (1.5-fold increase) in the proportion of IFN-γ producing CD8^+^ T cells post stimulation (Figure 3C), this effect was statistically significant only in the LAIV/Spn+ group due to the lower variance within the group (Figure 3C). TIV and control groups had overall no increase in the proportion of IFN-γ producing CD8^+^ T post stimulation with influenza antigens. In addition, IFN-γ production by lung TRM CD8^+^ T cells was not significantly altered post stimulation in any of the groups (Figure 3D).

Stimulation did not elicit production of IL-17A producing CD8^+^ T cells, except for IL-17A production by TRM CD8^+^ T cells in the Spn colonized group (2.6-fold increase, p = 0.008) (Supplementary Fig. 5C-D).

### LAIV increases frequency of influenza-specific IFN-γ producing TCR-γδ^+^ in the lungs of non-colonized individuals

TCR-γδ cells, a subset of specialized innate-like T cells that can exert effector functions immediately upon activation, play an important role in pulmonary infection^30, 31^. Therefore, we assessed whether TCR-γδ responses to influenza antigens were induced following vaccination. Although no significant increase in TNF-α producing TCR-γδ^+^ was observed in any of the groups (Figure 4A), the proportion of IFN-γ producing TCR-γδ^+^ was significantly greater in LAIV/Spn- group (median 2.9- fold increase upon stimulation compared to the unstimulated condition, p=0.002, Figure 4B). None of the other vaccinated or unvaccinated groups showed a significant induction of IFN-γ production. Similar to the other T cell subsets, IL-17A producing TCR-γδ^+^ cells did not significantly increase after stimulation with influenza antigens (Figure 4C).

**Figure 4.**
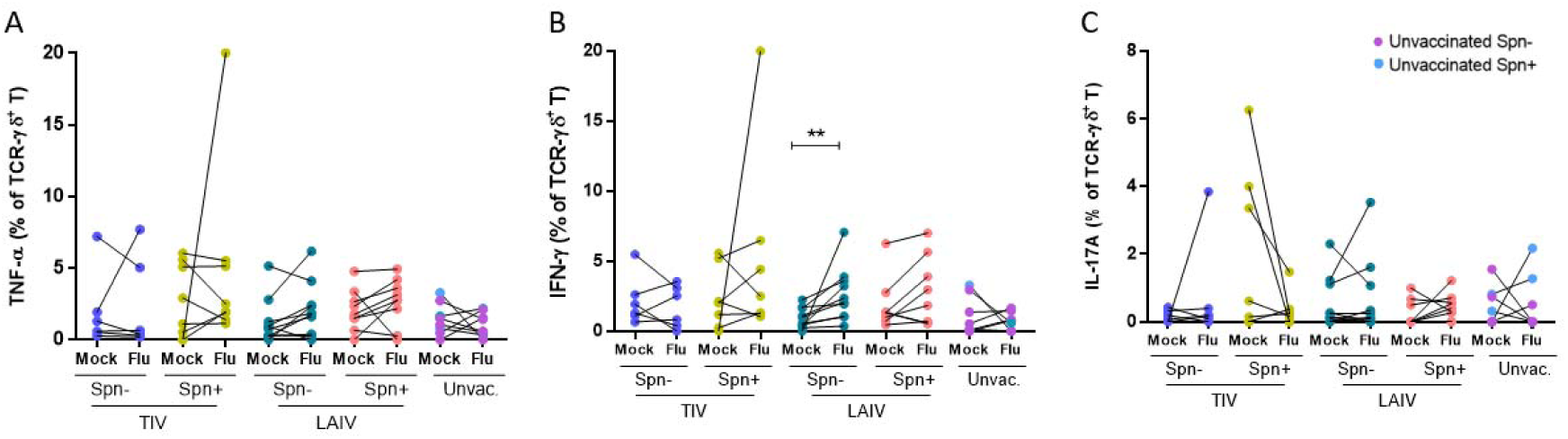
LAIV increases frequency of IFN-γ producing influenza-specific TCR-γδ^+^ in the lungs of Spn non-colonized individuals. Frequency of cytokine-producing TCR-γδ^+^ T cells was measured in human BAL samples by intracellular staining flow cytometry analysis after *in vitro* stimulation with influenza antigens or non-stimulation (mock). Volunteers were divided by vaccine and colonization status in TIV/Spn- (n=6), TIV/Spn+ (n=8), LAIV/Spn- (n=10), LAIV/Spn+ (n=9) and unvaccinated (n=8, 3 Spn- and 5 Spn+) group. Production of (A) TNF-α, (B) IFN-γ and **(C)** IL-17A by lung TCR-γδ T cells. Individual dot represents a single volunteer and the conditions per individual are connected. **p < 0.01 by Wilcoxon test.

### LAIV increases frequency of CD4^+^ regulatory T cells in the lung of non-colonized individuals

A balanced immune response in the lung has been demonstrated to be important in preventing pneumonia^32^. To investigate whether LAIV could alter frequency of regulatory T cells (T-regs) in the lung, we measured the frequency of CD25^hi^ FOXP3^+^ T-regs among CD4^+^ T cells using intracellular staining (Supplementary Fig. 2). Increased levels of CD4^+^ T- regs were only detected in BAL samples of LAIV/Spn- compared to unvaccinated individuals (mean 1.5-fold increase) (p = 0.039) (Figure 5).

**Figure 5.**
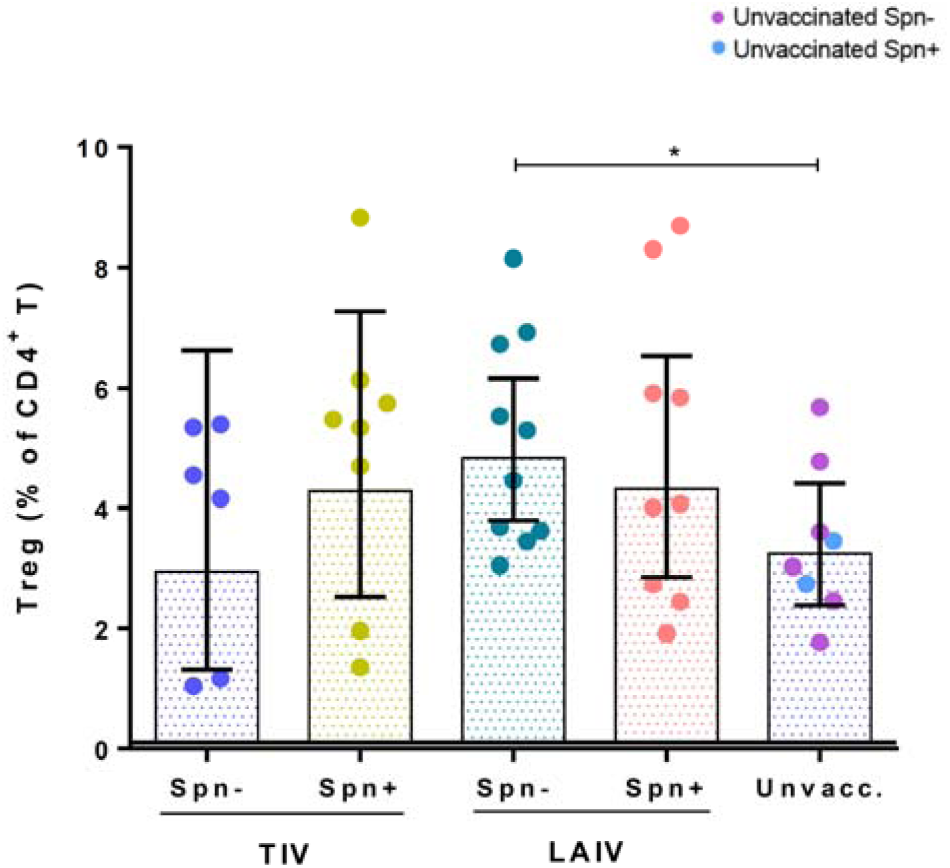
LAIV increases frequency of CD4^+^ regulatory T-cells in the lung of Spn non- colonized individuals. Frequency of unstimulated CD4^+^ T-regs (CD3^+^ CD4^+^ CD25^+^ FOXP3^+^) was measured by flow cytometry in human BAL samples from TIV/Spn- (n=6), TIV/Spn- (n=8), LAIV/Spn- (n=10), LAIV/Spn+ (n=9) and unvaccinated (n=8, 3 Spn- and 5 Spn+). Each individual dot represents a single volunteer and geometric means with 95% CI are shown. *p < 0.05 by unpaired t test.

### TIV but not LAIV vaccination increases levels of IgG to influenza in serum

In addition to cellular responses, we sought to assess humoral responses elicited by TIV and LAIV vaccination both systemically and at the mucosal sites (nasal and lung). In serum samples, IgG levels against influenza antigens were measured at baseline (prior to bacterial challenge and influenza immunization) and at Day 24 post vaccination. TIV induced a 5.9- fold increase (p < 0.0001) of influenza-specific IgG, while LAIV intranasal administration did not confer increase of sera IgG levels (Figure 6A). Prior colonization of the nasopharynx with Spn did not alter influenza-specific IgG levels induced in response to either vaccine. (Figure 6B).

**Figure 6.**
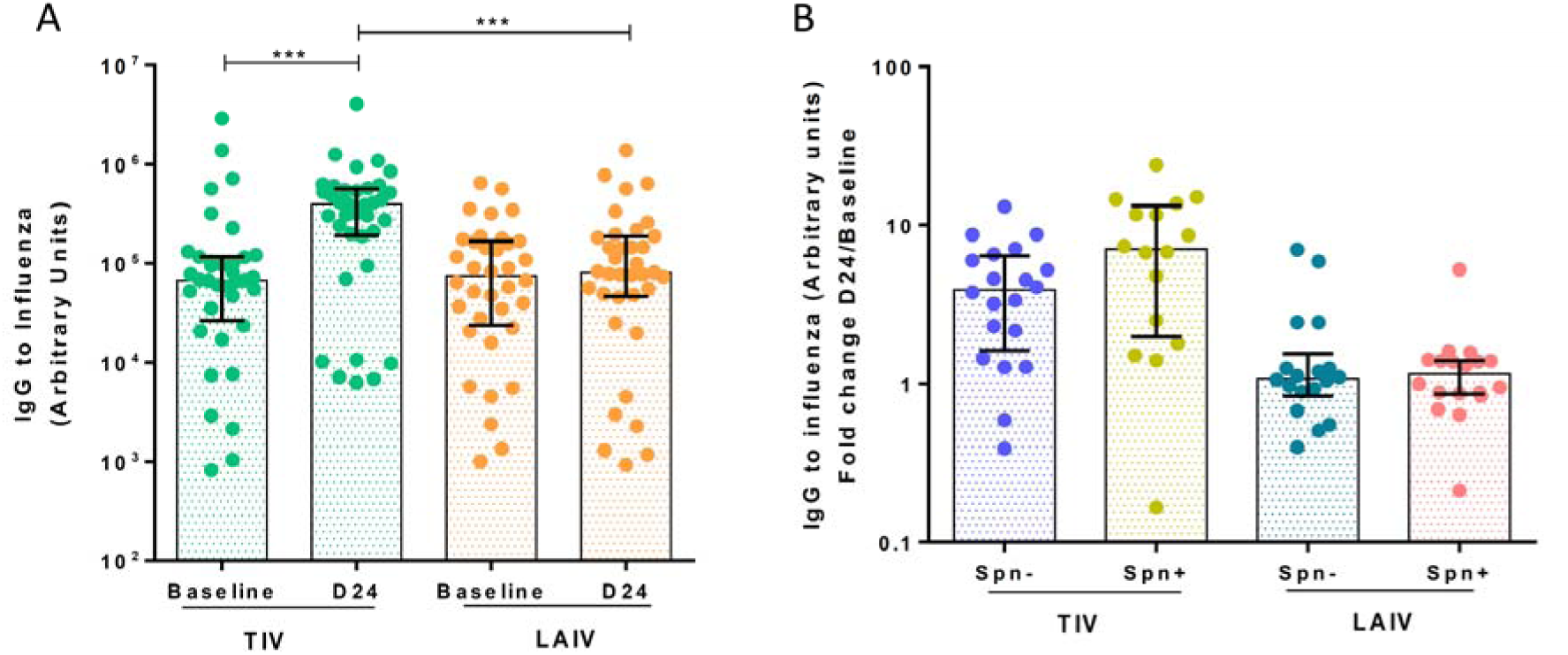
TIV but not LAIV vaccination increases levels of IgG to influenza in serum. **(A)** Geometric mean of IgG levels to influenza, measured by ELISA, in serum of LAIV (*n=36*) and TIV (*n=36*) vaccinated subjects at baseline (8 days pre-vaccination) and D24 (24 days post-vaccination). **(B)** Fold change (D24/Baseline) of paired IgG titres to influenza in serum following TIV or LAIV vaccination. TIV/Spn- (TIV vaccinated, but non-colonized with Spn, *n=20*), TIV/Spn+ (TIV vaccinated while Spn colonized, *n=16*), LAIV/Spn- (LAIV vaccinated but non-colonized with Spn, *n=18*); LAIV/Spn+ (LAIV vaccinated while Spn colonized, *n=18*). Medians with IQR are shown. ***p < 0.001 by Wilcoxon test for comparisons within the same group, and by Mann-Whitney test for comparisons between groups.

### *Streptococcus pneumoniae* colonization impairs nasal IgA induction following LAIV but does not alter responses to TIV

To assess antibody responses at the nasal mucosa, we measured influenza-specific IgA and IgG levels in nasal wash samples at baseline and 24 days following influenza immunization. TIV induced a median 2.2- and a 5.2-fold increase in influenza virus-specific IgA and IgG levels, respectively, 24 days post-vaccination (Figure 7A-B). LAIV also elicited an IgA and IgG antibody response, though both (IgA median 1.3-fold increase and IgG median 1.4-fold increase) were lower compared to those induced by TIV (Figure 7).

**Figure 7.**
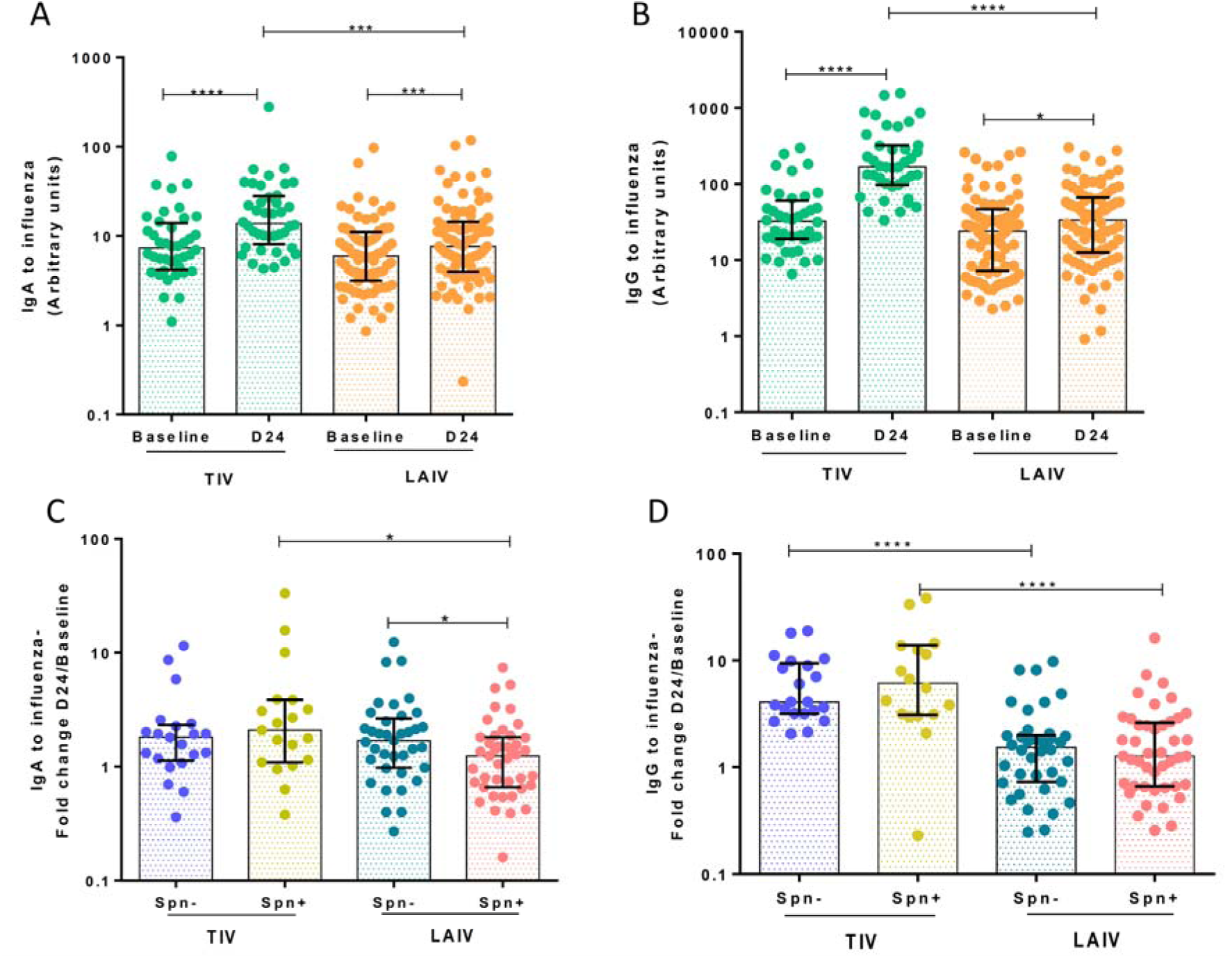
*Streptococcus pneumoniae* colonization impairs nasal IgA induction following LAIV but does not alter responses to TIV. **(A)** Geometric mean of IgA and **(B)** IgG titres to influenza measured by ELISA in nasal wash of TIV (*n=40*) and LAIV (*n=80*) vaccinated subjects at baseline (8 days pre-vaccination) and D24 (24 days post- vaccination). **(C)** Fold change (D24/Baseline) of paired IgA and **(D)** IgG titres to influenza in nasal wash following vaccination with TIV/Spn- (*n=21*), TIV/Spn+ (*n=19*), LAIV/Spn- (*n=37*), LAIV/Spn+ (*n=43*). Medians with IQR are shown. *p <0.05, ***p < 0.001, ****p < 0.0001 by Wilcoxon test for comparisons within the same group and by Mann-Whitney test for comparisons between groups.

LAIV-mediated immunogenicity at the nasal mucosa was also dependent on Spn colonization, as observed for the lung cellular responses. Pre-existing colonization of the nasopharynx with Spn affected IgA titres, but not IgG, in the LAIV group (Figure 7C-D). At day 24 post-vaccination, the LAIV/Spn- group had significantly greater levels of IgA to influenza circulating in the nasal lumen, compared to the LAIV/Spn+ group (LAIV/Spn- median= 1.69, IQR: 0.98-2.65 vs LAIV/Spn+ median=1 .24, IQR: 0.66-1.81) (p=0.02) (Figure 8C). Spn colonization did not alter antibody responses to influenza in the TIV group (Figure 7C-D).

**Figure 8.**
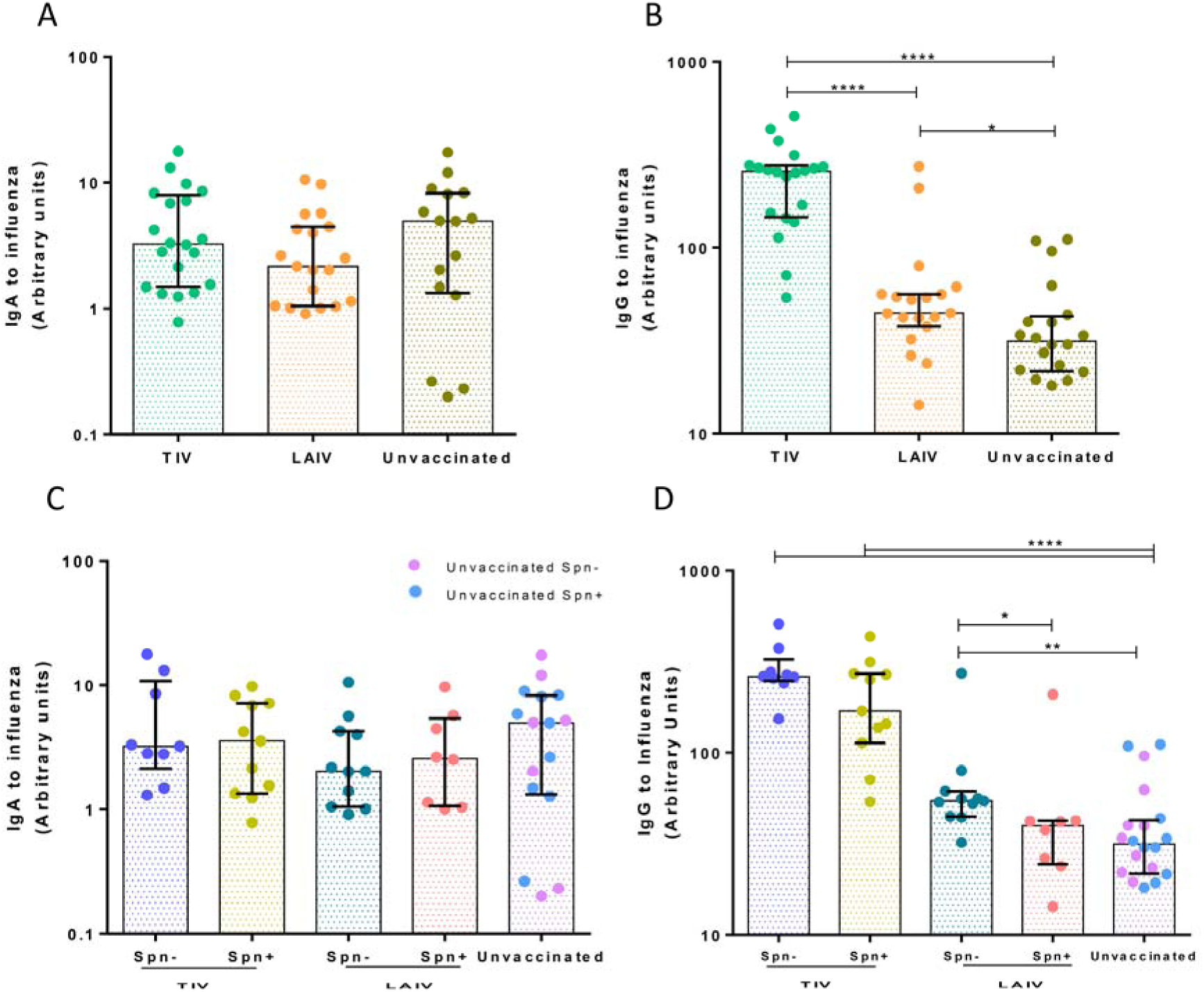
IgG but not IgA is induced by influenza vaccines in the lung, with LAIV responses being reduced during Spn colonization. **(A)-(B)** Geometric mean of IgA and IgG titres to influenza for TIV (n=20), LAIV (n=19) vaccinated subjects and unvaccinated (n=20) was measured by ELISA in BAL fluid. **(C)-(D)** Geometric mean of IgA and IgG titres grouped based on vaccination and colonization status, as TIV/Spn- (*n=9*), TIV/Spn+ (*n=11*), LAIV/Spn- (*n=11*), LAIV/Spn+ (*n=8*), unvaccinated (n=20). * p <0.05, ** p<0.01, **** p < 0.0001 by Wilcoxon test for comparisons within the same group and by Mann-Whitney test for comparisons between groups.

### IgG but not IgA is induced by influenza vaccines in the lung, with LAIV-mediated responses being impaired by pneumococcal colonization

Humoral responses in the lung following TIV or LAIV vaccination were assessed in bronchoalveolar lavage (BAL) samples collected between 26 to 46 days post influenza vaccination (Figure 8A). Due to the single timepoint sampling of the lung, 20 unvaccinated subjects (10 Spn-colonized and 10 non-colonized) were used as a control group.

Levels of IgA to influenza in the lung did not differ between TIV, LAIV and control groups (Figure 8A). In terms of IgG levels, TIV induced a high IgG response (median 5.8-fold increase compared to control) (p < 0.0001), whereas LAIV conferred a modest IgG induction (median 1.6-fold change compared to control) (p=0.028) (Figure 8B). TIV elicited influenza-specific IgG levels were 3.7x greater than LAIV-induced responses in the pulmonary mucosa (Figure 9B).

**Figure 9.**
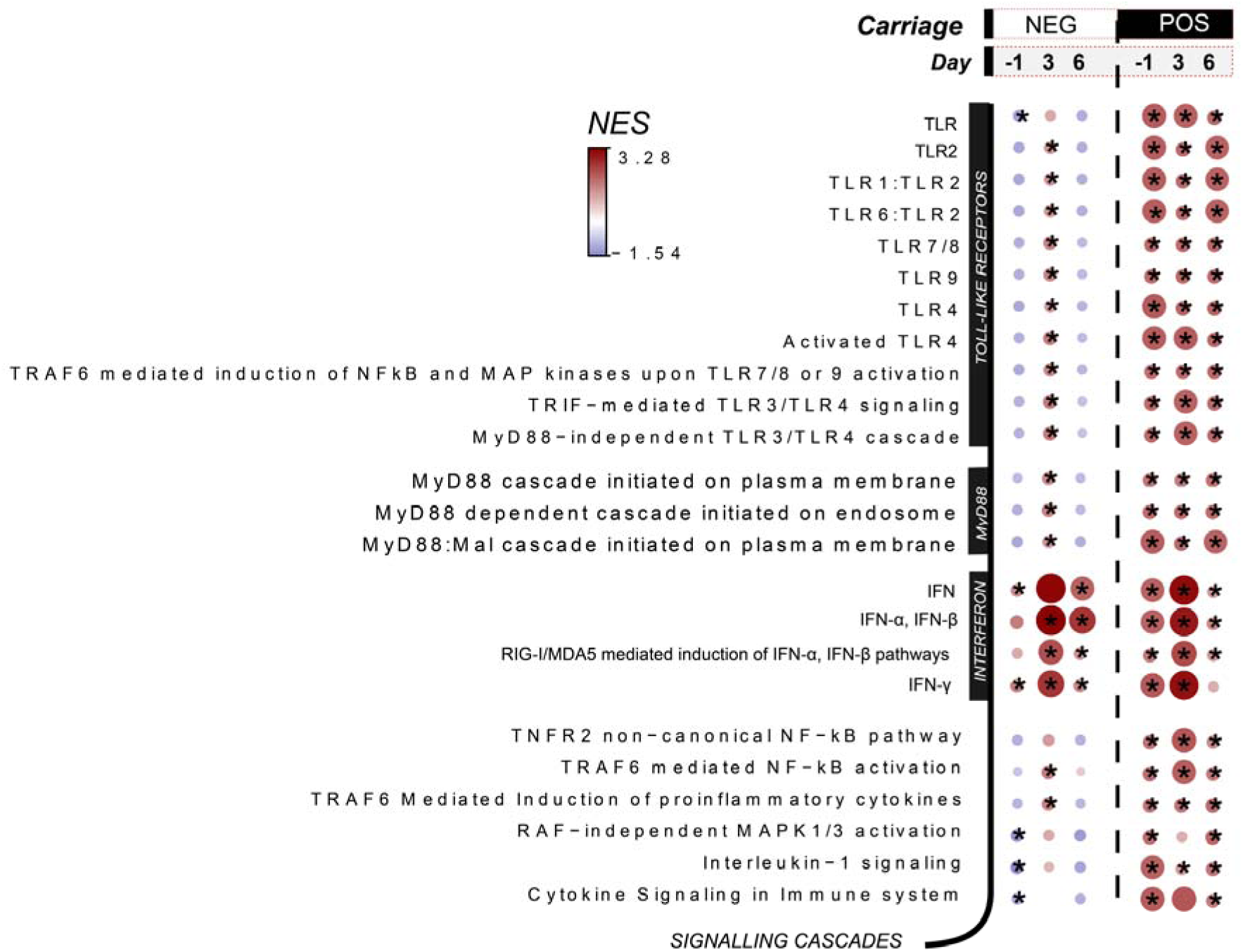
TLR priming by *S. pneumoniae* and increased type I IFN gene expression profile early post nasal colonisation. Selected pathways after Gene Set Enrichment Analysis (GSEA) for LAIV/Spn- (n=11) and LAIV/Spn+ (n= 9) groups at D-1, D3 and D6 in relation to LAIV administration applied on log2-fold changes (baseline/pre-Spn inoculation-normalised values). Normalised Enrichment Score (NES) is presented in gradient colour. Red shades indicate pathways over-presented, whereas blue shades depict the under-presented pathways at each time point in relation to baseline (prior pneumococcal inoculation). *p<0.05 by Wilcoxon paired test corrected by multiple-comparison testing (Benjamini-Hochberg).

IgA levels were not significantly increased in the lung by vaccination and therefore not affected by Spn colonization (Figure 8C). However, Spn colonization affected IgG titres in the LAIV vaccinated group, but not in the TIV group. IgG to influenza was higher in LAIV/Spn-group compared to the control group (1.73-fold increase (p=0.006), whereas the LAIV/Spn+ did not differ from the control group (Figure 8D).

### TLR priming by *S. pneumoniae* and increased type I IFN gene expression profile early post nasal colonisation establishment

To identify molecular signatures associated with reduced LAIV-mediated immunogenicity and impaired inflammatory responses due to established pneumococcal nasopharyngeal colonisation, we performed host RNA-sequencing on nasal cells at baseline, day −1 (before vaccination) and at days 3 and 6 post vaccination. Two days after the bacterial challenge but prior influenza vaccination, the LAIV/Spn+ group showed enrichment in genes related with TLR signalling, including TLR2, TLR4 and TLR9 (Figure 9). As expected, gene enrichment in TRL signalling was not observed in the LAIV/Spn-group at the same time point post inoculation. Additionally, the LAIV/Spn+ group exhibited enrichment in genes of interferon α/β, interferon-γ signalling and RIG-I/MDA5 mediated induction of IFN-α/β pathways (Figure 9). The upregulation of these pathways suggests that pneumococcal colonised volunteers had increased antiviral responses the day before the LAIV administration; a molecular profile that is likely to interfere with the influenza virus replication cycle in the nasopharynx.

To further investigate this observation, influenza RNA was quantified in unconcentrated nasal washes collected at Day 3 post vaccination in the LAIV group. Only ¼ of the LAIV vaccinated group had detectable levels of influenza viral RNA in the nose 3 days post vaccination, with no statistically significant differences in the levels of influenza viral RNA (Supplementary Fig. 6A) or in the percentage of shedders (CT < 40) between the Spn colonised (23.1%, 9/39) and non-colonised (27.5%, 11/40) (Supplementary Table 1). As expected, levels of influenza-specific IgA, following LAIV vaccination, were greater (2.5-fold change) in the nasal mucosa of volunteers with detectable viral influenza replication (Supplementary Fig. 6B). In contrast, raised influenza-specific IgG levels following vaccination did not differ between shedders and non-shedders (Supplementary Fig. 6C).

## DISCUSSION

We investigated the cellular and humoral immune responses elicited by TIV and LAIV, focusing on respiratory mucosa, and assessed whether colonization of the nasopharynx with *S. pneumoniae* influences vaccine immunogenicity. In agreement with previous studies^3^, TIV vaccination induced high systemic and mucosal antibody responses, whereas LAIV elicited both mucosal (mainly IgA) influenza virus-specific antibodies and cell-mediated immune responses. Interestingly, pre-existing pneumococcal colonization of the nasopharynx impaired host immunity to LAIV but did not alter TIV-induced responses. Antecedent pneumococcal colonization was also associated with weakened acute nasal pro-inflammatory responses post LAIV vaccination.

In the lungs, LAIV-induced cellular responses were heightened and significantly increased from those induced by TIV. LAIV nasal administration led to increased levels of TNF-α and IFN-γ producing CD4^+^ T cells, including TRMs, as well as TNF-α producing CD8^+^ T cells, upon *in vitro* stimulation. Interestingly, we observed that influenza specific CD4^+^ T cell lung responses were more pronounced in individuals not colonized with Spn at the time of vaccination suggesting increased immunogenicity of LAIV in the absence of pneumococcal colonization. Similarly, there was a higher proportion of IFN-γ producing TCR-γδ^+^ T cells in the non-colonized LAIV recipients. Moreover, LAIV was associated with increased frequencies of lung regulatory T cells, but only in the absence of nasal Spn colonization.

Humoral responses were highly induced by TIV, whereas LAIV conferred an overall modest antibody induction. Systemically, TIV elicited influenza virus-specific IgG responses, which were not observed in the LAIV vaccinated arm. In the nose, TIV conferred predominantly IgG induction, while LAIV was mainly associated with high levels of IgA. Colonization of the nasopharynx with *S. pneumoniae* at the time of LAIV administration impaired the induction of mucosal IgA to influenza in the nose and IgG in the lung. The modulatory effect of *S. pneumoniae* on adaptive immune responses to influenza virus has been previously reported in a murine co-infection model, highlighting the importance of the history of pathogen exposures, which can critically affect the generation of protective antiviral antibodies and subsequently reduce influenza vaccination efficacy^23^.

The protection provided by LAIV relies on a transient viral replication in the nasopharynx in order to induce sufficient antibody levels to influenza, which we observed with increased IgA induction in shedders compared to non-shedders. LAIV in adults, unlike children, does not confer superior protection compared to TIV^33^. An explanation consistent with the hypothesis is that life-long accumulation of influenza immunity through natural exposure and previous vaccinations can prevent the nasal replication of the attenuated virus and shorten the viral replication cycle^34^. Here, 25.3% of young adults vaccinated with LAIV shed attenuated influenza virus (either influenza A or B), in contrast to the much higher shedding rates observed in 2 to 5 year olds children in other studies^35^. Taking under consideration virus neutralization by pre-existing antibodies against influenza due to life-time exposure or a shortened virus replication cycle^34^, increased influenza shedding had to has been detected in the LAIV cohort in the first two days after the vaccine administration. Consequently, LAIV may elicit less potent responses in adults compared to children, thus any extrapolation from findings in adults to children, the target population for this vaccine, must be done with caution.

Children display high rates of Spn colonization^36, 37^. Our finding that concurrent Spn colonization could inhibit LAIV-induced immune responses, is another variable that should be taken into account when evaluating LAIV efficacy. This phenomenon could explain the lack of efficacy reported in Senegal^38^, as Spn colonization rates are higher in low-income countries^39^. The impaired LAIV-induced immunity during established Spn colonization was associated with a lack of a pro-inflammatory response in the nasal mucosa following LAIV vaccination. An explanation for this is that Spn colonization affects local immune and epithelial cell responses upon LAIV vaccination, which could diminish immune cells infiltration and antigen presenting cells (APC) activation, impacting on the downstream memory responses^40, 41^. It is also possible that Spn colonization interferes with the viral replication cycle ^42, 43^, through stimulation of TLR signalling. A number of studies have reported the broad contribution of TLR2 to the antiviral interferon response by indirectly governing the production of IFNs induced by other Toll like receptors, as wells as downstream of the cytosolic Rig-I like receptors^44, 45^. In an infant mouse influenza A - *S. pneumoniae* co-infection model, mice deficient for TLR2 showed decreased expression of IFN-α and higher viral titres than the wildtype animals, with this great viral burden to correlate with heightened inflammation^46^. In our study, Spn colonised volunteers upregulated genes involved in TLR2 signalling, RIG-I/MDA5 mediated induction of IFN-α/β and IFN-α/β pathways before exposure to LAIV and exhibited impaired inflammatory responses post vaccination. Despite these observations, any alteration of viral replication cycle mediated by Spn colonisation was limited by the late time point of viral quantification at day 3 post LAIV administration. An alternative hypothesis of the curtailed viral shedding in the LAIV/Spn+ group would be the inhibitory effect of pneumococcal neuraminidases, particularly NanA, on influenza virus attachment to the epithelium, as shown in an infant mouse model of S. pneumoniae-IAV coinfection model^47^. In light of these observations, it would also be interesting to investigate to what extent symptoms and inflammation caused by wild type influenza viruses are altered by concurrent Spn colonization in humans.

Ideally, an effective and broadly protective influenza vaccine should induce both humoral and cellular immunity. Whereas antibody responses to influenza show some degree of strain cross-reactivity^48, 49^, they are insufficient to provide heterosubtypic, cross-strain influenza protection^50, 51^. Recent data from longitudinal cohort studies of naturally acquired infection have highlighted the potential of T-cells as key players in mediating heterosubtypic immunity in humans^52, 53^. We observed that even in the absence of vaccination, healthy adults showed CD4^+^ T cell responses to influenza stimulation, which likely reflects their lifelong exposure to influenza viruses. The use of purified, adjuvanted influenza antigens (TIV) as the stimulus to measure cellular responses *in vitro*, would possibly lead to greater T cells responses. Our results demonstrated that LAIV induced influenza-specific cytokine-producing CD8^+^ and CD4^+^ T-cells, including TRM in the lung. Such cells are important during influenza infection in protection of mucosal barrier tissues against pathogen challenge by producing chemokines for cell recruitment^54^. It has been shown that TRM T cells provide superior protection to influenza infection when compared with circulating T cells^55^. By seeding the lungs with these cells, it is possible to establish long-term heterosubtypic protection to influenza ^56, 57^.

We have also demonstrated that, in volunteers not colonized by Spn, LAIV increased levels of Tregs in the lung compared to unvaccinated individuals. CD4^+^ Tregs contribute to homeostasis of the immune system, controlling infection by respiratory viruses and avoiding secondary bacterial infection^58^. As a result of recurrent exposure to virus and bacteria, CD4^+^ Tregs increase in frequency with age^59^. For this reason, our findings in adults might underestimate the effect of LAIV on frequency of Tregs in the lung of children.

Although, LAIV and TIV mediated responses were assessed in the context of a randomised clinical trial, the study was limited by evaluation of a single pneumococcal serotype in healthy adults-likely to have neutralizing influenza antibodies. Any LAIV effect in children may be differential due to lower antibody titres or higher natural rates of colonization.

In conclusion, using a controlled human model in which pneumococcal infection occurred at a known time relative to vaccination, we were able to highlight differences in immunogenicity between LAIV and TIV at relevant mucosal sites. Moreover, we identified *S. pneumoniae* colonization as an important variable in LAIV-induced immunity.

## METHODS

### Study Design

Adult volunteers were enrolled in the parent LAIV clinical trial study (REC 14/NW/1460)^26^. Exclusion criteria included: a prior history of influenza or pneumococcal vaccination; clinically confirmed pneumococcal disease in the preceding two years; pregnancy; close contact with individuals at increased risk for pneumococcal disease (children under 5, immunosuppressed, elderly); recent febrile illness; current or recent use of antibiotics or immune-modulating medication. Participants were inoculated with 80,000 CFU per nostril of serotype 6B as previously described ^25^. All volunteers received influenza vaccination 3 days post pneumococcal inoculation. LAIV group (n=80) received the live attenuated influenza vaccine (2016/2017 Fluenz Tetra, AstraZeneca, UK), whereas TIV group (n=90) received the tetravalent influenza vaccine, (2016/2017 Fluarix Tetra, GlaxoSmithKline, UK). Overall rates of carriage were not different between LAIV and control groups by conventional microbiology (37/80 [46.3%] vs 45/90 [50.0%] respectively.

For investigation of immune responses, samples of nasal wash, nasal lining fluid, nasal cells, bronchoalveolar lavage, serum were collected from volunteers at specific timepoints, processed and frozen for future analysis (Supplementary Figure 1). For comparisons within the lung datasets, BAL fluid and lung lymphocytes from an unvaccinated EHPC group (n=20, 10 Spn- and 10 Spn+) were used as a control.

### Ethics Statement

Ethical approval was given by NHS Liverpool East Research Ethics Committee (REC), reference number 14/NW/1460. The trial was registered on EudraCT, Protocol 2014- 004634-26 (NCT ID: NCT03502291). All volunteers gave written informed consent and research was conducted in compliance with all relevant ethical regulations. BAL samples of the control (non-vaccinated cohort) were collected as part of separate EHPC clinical trial (NHS North-West REC, reference 15/NW/0931).

### Detection of Spn colonization

To detect bacteria in the nasopharynx, nasal wash samples plates were examined by classical microbiology for presence of Spn as previously described^60, 61^. Colonized individuals were defined as anyone who had a positive nasal wash sample at any timepoint following inoculation.

### Bronchoalveolar lavage (BAL) analysis

Bronchoscopy was performed using topical anaesthesia and BAL was collected as described previously ^65^. Briefly, a total of 200 mL of warm 0.9% saline was instilled and retrieved from a sub-segmental bronchus of the right middle lobe by hand suction. The BAL was placed into sterile tubes on ice. BAL was processed as described^62^. In short, the BAL sample was filtered to remove mucus and centrifuged at 400g for 10 minutes. BAL cells were re-suspended in RPMI with antibiotic mixture (Penicillin-Streptomycin-Neomycin, Thermo-Fisher, Waltham, MA, USA). Cells were plated in a 24-well plate (Greiner Bio-One, Kremsmünster, Austria) to allow macrophages to adhere for 4 hours at 37°C, 5% CO_2_. Non-adherent BAL cells were collected, washed, centrifuged at 200g for 10min and resuspended in PRMI prior stimulation.

### Intracellular cytokine staining

Non-adherent BAL cells were counted and incubated at 1×10^6^ cells/mL in medium with RPMI, FBS (10% heat inactivated, Thermo-Fisher) and antibiotic mixture (Penicillin-Streptomycin-Neomycin) at 37°C. Samples were stimulated with 1.2 μg/mL influenza antigens (TIV, tetravalent influenza vaccine, 2016/2017) or left unstimulated as negative control, and incubated for 2 hours. Then, 1000x diluted BD Golgiplug (BD Biosciences, San Jose, California, USA) was added and cells were cultured for an additional 16 hours as previously described ^66^.

After 16 hours, the cells were washed with 3 mL of PBS, resuspended and stained with Violet Viability dye (LIVE/DEAD Fixable Dead Cell Stain kit, Invitrogen, UK). After 15 minutes, the cells were stained with the surface markers CD3-APCH7 (clone SK7), TCR-γδ– PECy7 (clone 11F2) from BD Biosciences (San Jose, California, USA), CD4–PerCP5.5 (clone SK3), CD8–AF700 (clone SK1), CD69–BV650 (clone FN5O), CD25-PE.TxsRed (clone M-A251), CD103–BV605 (clone Ber-ACT8), CD49a-APC (clone TS2/7) from Biolegend (San Diego, CA) and incubated for 15 minutes. Cells were fixed and permeabilized using the Foxp3/Transcription Factor Staining Buffer Set (eBiosciences, San Diego, CA) as per manufacturer’s instructions. Cells were then stained with intracellular markers FOXP3-FITC (clone 259D), IFN-γ-PE (clone 4S.B3), TNF-α–BV711 (cloneMAb11) Biolegend (San Diego, CA) and IL-10–BV786 (clone JES3-9D7) IL-17A–BV510 (clone N49- 653) from BD Biosciences (San Jose, California, USA). After 30 minutes, samples were washed with 3 mL of PBS and resuspended in 200 µL of PBS and acquired on a BD LSR flow cytometer (Becton Dickinson, UK). Flow cytometry data was analysed using FlowJo cell analysis software version 10 (FlowJo, LLC, Ashland, Ore).

### Quantitative reverse transcription-PCR (qRT-PCR)

QRT-PCR was used to quantify nasal virus shedding on volunteers vaccinated with LAIV. RNA was isolated (RNeasykit; Qiagen) from nasal wash fluid, following generation of cDNA (high-capacity RT kit; Applied Biosystems) for use in quantitative PCR (SYBR Green PCR master mix; Applied Biosystems). Samples were tested using primers, probes and PCR assay conditions specific for human influenza virus A and B^63^. Results were analysed using the threshold cycle (2⍰ΔΔCT) method by comparison to GAPDH (glyceraldehyde-3- phosphate dehydrogenase) transcription.

### Enzyme-linked immunosorbent assay (ELISA)

ELISA was used to quantify levels of IgG and IgA antibodies to influenza in the serum, nasal wash and BAL supernatant of volunteers vaccinated with TIV or LAIV. Pooled sera of 7 TIV vaccinated volunteers was heat-inactivated (at 56°C for 30 min) and used as standard in both total IgA and IgG to influenza ELISA. Antibody levels were expressed in arbitrary units relative to this standard curve. For IgG detection, an initial standard dilution of 1:4000 was used, while for IgA it was diluted 1:40.

Briefly, 96-well plates (Nunc, Denmark) were coated with 100 µL of 0.2 µg/mL TIV in PBS overnight at room temperature. After the overnight incubation, plates were washed following blocking with 100 µL of PBS with 1% bovine serum albumin (BSA) for one hour at room temperature. Then, plates were washed, and samples were added in duplicates and incubated for 2 more hours at room temperature. Each wash consisted of washing the plate three times with PBS with 0.005% Tween 20 (Sigma, Germany).

For detection of IgG and IgA, a 1:5000 and 1:4000 dilution of anti-human-IgG (Sigma, A9544, Germany) and anti-human-IgA (Sigma, A9669, Germany), respectively, was made using 0.1% BSA and 100 µL added to each well after washing and incubated at room temperature for 2 hours.

Then, for both IgA and IgG to influenza, plates were washed and 100 µL of p-Nitrophenyl Phosphate (Sigma-Aldrich, Poole, U.K.) was added to the wells. The optical density of each well was measured at 405 nm using a FLUOstar Omega ELISA microplate reader (BMG Labtech), the average blank corrected value was calculated for each sample and the data analysed using Omega Analysis (BMG Labtech).

### Luminex analysis of nasal lining fluid

Nasal lining fluid was collected using nasosorption filters as previously described^64^ and stored at −80°C until analysis. Prior to analysis, cytokines were eluted from stored filters using 100μl of assay diluent buffer (ThermoFisher) by centrifugation. The eluate was cleared by further centrifugation at 16,000g for 10 min. Samples were and acquired on a LX200 using a 30-plex magnetic human Luminex cytokine kit (ThermoFisher) and results were analysed with xPonent3.1 software following manufacturer’s instructions. Samples were analysed in duplicates and cytokines with a CV>25% for a given sample were excluded from further analysis.

### RNA extraction and sequencing

Nasal cells were collected in RNALater (ThermoFisher) at −80°C until extraction. RNA extraction was performed using the RNEasy micro kit (Qiagen) with on column DNA digestion. Extracted RNA was quantified using a Qubit™ (ThermoFisher). Sample integrity assessment (Bioanalyzer, Agilent), library preparation and RNA-sequencing (Illumina Hiseq4000, 20M reads, 100 paired-end reads) were performed at the Beijing Genome Institute (China).

### RNA sequencing analysis

Quality control of raw sequencing data was done using fastQC. Mapping to a human reference genome assembly (GRCh38) was done using STAR 2.5.0a^65^. Read counts from the resulting BAM alignment files were obtained with featureCounts using a GTF gene annotation from the Ensembl database^66, 67^. The R/Bioconductor package DESeq2 was used to identify differentially expressed genes among the samples, after removing absent features (zero counts)^68^. Genes with an FDR value < 0.1 and an absolute fold change (FC) > 1.5 (baseline-normalised values) were identified as differentially expressed. For each timepoint comparison, gene set enrichment analysis (GSEA) was performed using the fgsea R package (REF1). Genes with Ensembl IDs were transformed into Gene Symbols by the biomart package (REF2, REF3) and ordered by its log fold-change values. Pre-ranked genes and Reactome gene sets from Enrichr (REF4, REF5) were provided to fgsea, with remaining default parameters. To identify significant common pathways between all comparisons, pathways with a p-value below a threshold of 0.05 for at least one comparison were selected and clustered based on the Normalized Enrichment Scores (NES) with hierarchical clustering. Correlation plots were generated to display the NES values using the corrplot package (REF6).

### Statistical analysis

All sampling, processing and data analysis were performed while blinded to vaccination group to not bias results. Non-parametric tests were used for statistical analysis where number of samples were insufficient for a normal distribution of results. Statistics were calculated in GraphPad prism version 6.0 and 7.0 for Windows (GraphPad Software, California USA) and R Statistical Software (R Foundation for Statistical Computing). Differences were considered statically significant if p ≤ 0.05. Benjamini-Hochberg multiple correction was performed in R on both 30-plex cytokine data and RNAseq data analysis.

## Data Availability

Raw RNA-sequencing data will be deposited in the GEO repository
All datasets used and analysed during the current study will be available from the Liverpool school of Tropical Medicine online repository. This can be accessed via: http://archive.lstmed.ac.uk/.

## Acknowledgements

The authors thank the Data Monitoring and Safety Committee (Brian Faragher, Christopher Green, and Robert C. Read). We would like to thank all the volunteers for their participation in the clinical trial and the NWC CLRN for their continued support. Also, the clinical team members for helping to collect the samples, as well as Dr Dessi Loukov for helpful review of the manuscript. This work was funded by grants from BMGF (OPP1117728) awarded to DMF and MRC grants (MR/M011569/1) awarded to SBG. The funders had no role in study design, data collection and analysis, decision to publish, or preparation of the manuscript. Flow cytometric acquisition was performed on a BD LSRII funded by WelIcome Trust Multi-User Equipment grant (104936/Z/14/Z).

## Author Contributions

D.M.F., S.P.J. and E.M. conceived and designed the study. B.F.C., J.Reine, E.Negera, E.N., S.P. and D.B. acquired the data. B.F.C., F.M, J.Reine, E.N., H.N., S.P, H.N., D.M.F, S.P.J, and EM analysed and interpreted the data. J.R., S.Z., A.C., S.C. and V.C. assisted in clinical procedures and recruitment. B.F.C wrote the first draft of the paper. B.F.C., J.R., S.Z., J.Reine, E.Negera, E.N., S.P., C.S. A.C., V.C., D.B., S.B.G., H.N., D.M.F, S.P.J. and E.M. commented on and approved the paper.

